# Post-EVT CTP Imaging as a Patient-Selection Tool for Adjuvant Therapy: Review, Meta-Analysis, and Clinical Threshold Framework

**DOI:** 10.64898/2026.05.01.26352264

**Authors:** Roni Eichel, Michael Teitcher, Stefan Mausbach, Alona Poplavska, Salam Shqair, Roi Eichel, Eliel Ben David, Vladimir Borodetsky, Natan M Bornstein

## Abstract

**Background and Purpose:** Despite high rates of macrovascular recanalization, approximately half of patients with large vessel occlusion stroke fail to achieve functional independence after endovascular thrombectomy (EVT). Residual tissue-level perfusion abnormalities on post-procedural CT perfusion (CTP) may indicate futile recanalization and inform selection for adjuvant therapy. We synthesized post-EVT CTP thresholds, summarized acquisition timing, and discussed implications for patient selection in trials of intra-arterial thrombolysis, antithrombotics, and neuroprotection, limited to studies performing perfusion imaging after EVT.

**Methods:** We searched MEDLINE, EMBASE, and the Cochrane Library (January 2018–April 2026) for studies performing perfusion imaging after EVT, reporting ≥1 quantitative CTP parameter with functional or neurological outcome, and enrolling ≥10 patients; pre-EVT CTP studies were excluded. Functional independence with versus without post-EVT hypoperfusion was pooled using DerSimonian–Laird random-effects. Individual patient data from our prospective Cerebrolysin proof-of-concept cohort (N=18) were integrated.

**Results:** Nine post-EVT perfusion imaging studies (497 patients) met inclusion criteria. Residual hypoperfusion occurred in 21–53% of angiographically successful reperfusions and was associated with lower odds of functional independence (pooled OR 0.23, 95% CI 0.17–0.33; I²=29%). A Tmax >6 s volume <3.5 mL at 30–90 minutes post-EVT was the most consistently validated threshold (OR 3.5, 95% CI 1.6–7.8). In our cohort, an ischemic core (rCBF <30%) of 0 mL versus any detectable residual core was associated with markedly higher odds of independence (OR 27.5, 95% CI 1.0–746 with continuity correction; ρ=0.77, p=0.003). The optimal CTP acquisition window is 30–120 minutes post-EVT.

**Conclusions:** Post-EVT CTP outperforms modified TICI grading for predicting functional outcome and identifies biologically distinct subgroups for adjuvant therapy selection. Standardized post-EVT CTP at 30–120 minutes, applied with the proposed threshold framework, should be used for eligibility and stratification in future trials of intra-arterial thrombolysis, antithrombotics, and neuroprotection.

## Introduction

Endovascular thrombectomy (EVT) has fundamentally transformed the management of acute ischemic stroke due to large vessel occlusion (AIS-LVO). The HERMES collaboration, pooling individual patient data from five randomized trials (N=1287), demonstrated successful macrovascular recanalization in 71% of treated patients, with functional independence (modified Rankin Scale [mRS] 0–2 at 90 days) achieved in 46% of the EVT group versus 26% of controls.^1^ Despite this transformative efficacy, the persistent gap between angiographic success and clinical recovery remains one of the most pressing problems in stroke neurology: approximately half of all successfully recanalized patients fail to achieve independence, a phenomenon termed “futile recanalization” or “clinically ineffective reperfusion.”^2,3^

Multiple mechanisms have been proposed. The no-reflow phenomenon — defined as persistent microvascular hypoperfusion despite macrovascular patency — occurs in approximately 29% of patients with angiographically successful reperfusion.^4^ Procedural factors, particularly multiple thrombectomy passes, contribute to distal embolization and microvascular injury.^5^ Inadequate collateral circulation, early infarct core expansion, reperfusion injury, and failure of neurovascular unit integrity further compound the problem.^6,7^ Together, these mechanisms underscore that the modified Thrombolysis in Cerebral Infarction (mTICI) score, while clinically essential for procedural reporting, captures only macrovascular patency^7^ and provides an incomplete picture of tissue-level reperfusion.^8^

CT perfusion (CTP) imaging — used routinely before EVT to identify salvageable penumbra — has more recently attracted attention as a real-time post-procedural biomarker. Performed 30–120 minutes after thrombectomy, CTP delineates regions of persistent hypoperfusion (commonly quantified by the volume of tissue with time-to-maximum [Tmax] >6 seconds), residual ischemic core (relative cerebral blood flow [rCBF] <30%), complete reperfusion, and paradoxical hyperperfusion with spatial precision unavailable from angiography alone.^9^ Early prospective studies have demonstrated that post-EVT CTP hypoperfusion volume is a stronger predictor of final infarct volume and functional outcome than the mTICI score obtained at the end of the procedure, and may identify patients in whom delayed clinical recovery is plausible despite absence of early neurological improvement.^9,10^

The clinical importance of this biomarker is amplified by a rapidly expanding pipeline of post-EVT adjuvant therapies. Intra-arterial thrombolytics (alteplase, tenecteplase) have shown benefit when administered after incomplete macrovascular reperfusion.^11^ Antiplatelet escalation with glycoprotein IIb/IIIa inhibitors (tirofiban, eptifibatide) has been evaluated as an adjunct to EVT.^12^ Neurotrophic and pleotropic agents — most notably cerebrolysin, citicoline, and edaravone — represent the closest current candidates for true post-EVT pharmacological neuroprotection: while no agent has yet achieved regulatory approval as a post-EVT neuroprotectant, several have demonstrated safety and signals of benefit in stroke trials and are under prospective evaluation. Identifying the right patient for the right adjuvant therapy in real time is the central challenge, and post-EVT CTP is currently the best available imaging instrument for this task.^9,11^

Despite this rationale, the literature remains heterogeneous. A critical methodological confound has persisted in published reviews: several have included pre-EVT admission CTP data — which measure baseline ischemic burden and collateral status — alongside genuinely post-EVT perfusion data. This conflation produces threshold estimates that do not reflect the post-procedural reperfusion milieu and are not applicable to adjuvant therapy selection. The present systematic review and meta-analysis addresses this gap by restricting inclusion exclusively to studies performing perfusion imaging after EVT completion, and by integrating individual patient data from our prospective proof-of-concept cohort of 18 patients treated with intravenous cerebrolysin following EVT with residual perfusion deficits.

## Methods

### Search Strategy and Eligibility Criteria

This systematic review and meta-analysis were conducted and reported in accordance with the PRISMA 2020 statement.^13^ MEDLINE (via PubMed), EMBASE, and the Cochrane Library were searched from January 2018 through April 2026. The lower bound was selected to follow the publication of the DAWN and DEFUSE 3 trials, which established CT perfusion as standard practice for EVT eligibility assessment and reframed the role of perfusion imaging in the EVT workflow.^14^ Search terms were: (“endovascular thrombectomy” OR “mechanical thrombectomy” OR “endovascular treatment”) AND (“CT perfusion” OR “computed tomography perfusion” OR “perfusion imaging” OR “MRI perfusion” OR “magnetic resonance perfusion” OR “no-reflow”) AND (“post-procedural” OR “after thrombectomy” OR “post-thrombectomy” OR “post-recanalization” OR “residual hypoperfusion”). No language restrictions were applied. Reference lists of included studies and relevant systematic reviews were hand-searched.

Inclusion criteria were: (1) prospective or retrospective cohort study or registry analysis; (2) perfusion imaging performed after completion of EVT; (3) reporting of at least one quantitative post-EVT CTP parameter (Tmax volume, ischemic core volume, hypoperfusion intensity ratio [HIR], CBF/CBV maps, or microvascular perfusion classification); and (4) reporting of functional outcome (mRS) or neurological outcome (NIHSS).

Exclusion criteria were: (1) perfusion imaging performed exclusively before EVT; (2) experimental or preclinical studies; (3) case reports or series with fewer than 10 patients; (4) conference abstracts without full published data, with the sole exception of our own cohort for which complete individual patient data are available; and (5) systematic reviews or meta-analyses whose constituent studies overlap with individually-included primary studies (excluded from the study inventory; cited only as supporting contextual evidence). Studies measuring pre-EVT admission CTP parameters to predict outcomes or guide patient selection were explicitly excluded, even when post-EVT clinical outcome data were reported, as these do not measure post-procedural reperfusion status.

### Data Extraction

Two investigators (R.E. and N.M.B.) independently extracted from each included study: design (single-or multi-centre), sample size, demographics (age, sex, admission NIHSS), imaging modality and software platform, timing of post-EVT perfusion imaging relative to end of procedure, CTP parameters reported (Tmax >6 s volume, ischemic core rCBF <30%, hypoperfusion intensity ratio [HIR; defined as the volume of tissue with Tmax >10 s divided by the volume with Tmax >6 s], CBF/CBV microvascular patterns), final mTICI grade distribution, functional outcome (mRS 0–2 at 90 days or latest available follow-up), NIHSS at discharge, hemorrhagic transformation rates, and specific threshold values reported for predicting good versus poor outcomes. Discrepancies were resolved by consensus with a third investigator (S.M.).

### Statistical Analysis

For studies reporting dichotomized rates of functional independence (mRS 0–2) in patients with versus without post-EVT hypoperfusion, unadjusted odds ratios (OR) with 95% confidence intervals (CI) were calculated or directly extracted. Pooled ORs were estimated using the DerSimonian–Laird random-effects estimator. We selected DerSimonian–Laird in preference to restricted maximum likelihood (REML) given the small number of constituent studies (k=4 for the primary pooled analysis), in which the REML estimator can underestimate between-study variance and may suffer convergence problems; the DerSimonian–Laird method has well-characterized performance in this setting and is the most widely-used approach in published meta-analyses of similarly-sized neurovascular literature.^15^ Sensitivity analyses with the REML estimator yielded effect estimates that differed by less than 0.02 log-OR and did not alter the directional conclusions. Between-study heterogeneity was quantified using the I² statistic. CTP threshold values were synthesized qualitatively within three imaging timing sub-groups: immediate (<30 minutes post-EVT), early (30–120 minutes), and delayed (>2 hours). For our own cohort, where one cell of the 2×2 contingency table was zero, the Haldane–Anscombe continuity correction (adding 0.5 to all cells) was applied to compute the OR and 95% CI; the Spearman rank correlation between post-EVT ischemic core volume and mRS was computed in parallel.

## Results

### Study Selection and Characteristics

Database searches identified 847 unique records after deduplication. After title and abstract screening, 68 full-text articles were evaluated. Nine individual cohort studies met all inclusion criteria and were included in the primary study inventory. One systematic review and meta-analysis (Mujanovic et al., Int J Stroke 2024, N=719) was identified but excluded from the study inventory because it demonstrably subsumes individually-included primary studies — Rubiera 2020 is explicitly cited as reference 11 within that meta-analysis, confirming direct overlap. Including it as an independent table entry would constitute double-counting; it is therefore cited only as external contextual validation.^4^ The most common reasons for exclusion of full-text articles were use of exclusively pre-EVT CTP data (n=14), case series below the minimum sample size (n=6), purely qualitative perfusion assessment without quantitative parameters (n=4), and unavailable full-text data (n=3). The nine included cohort studies enrolled a total of 497 patients with non-overlapping populations. Characteristics of included studies are summarized in Table 1.

**Table 1.**
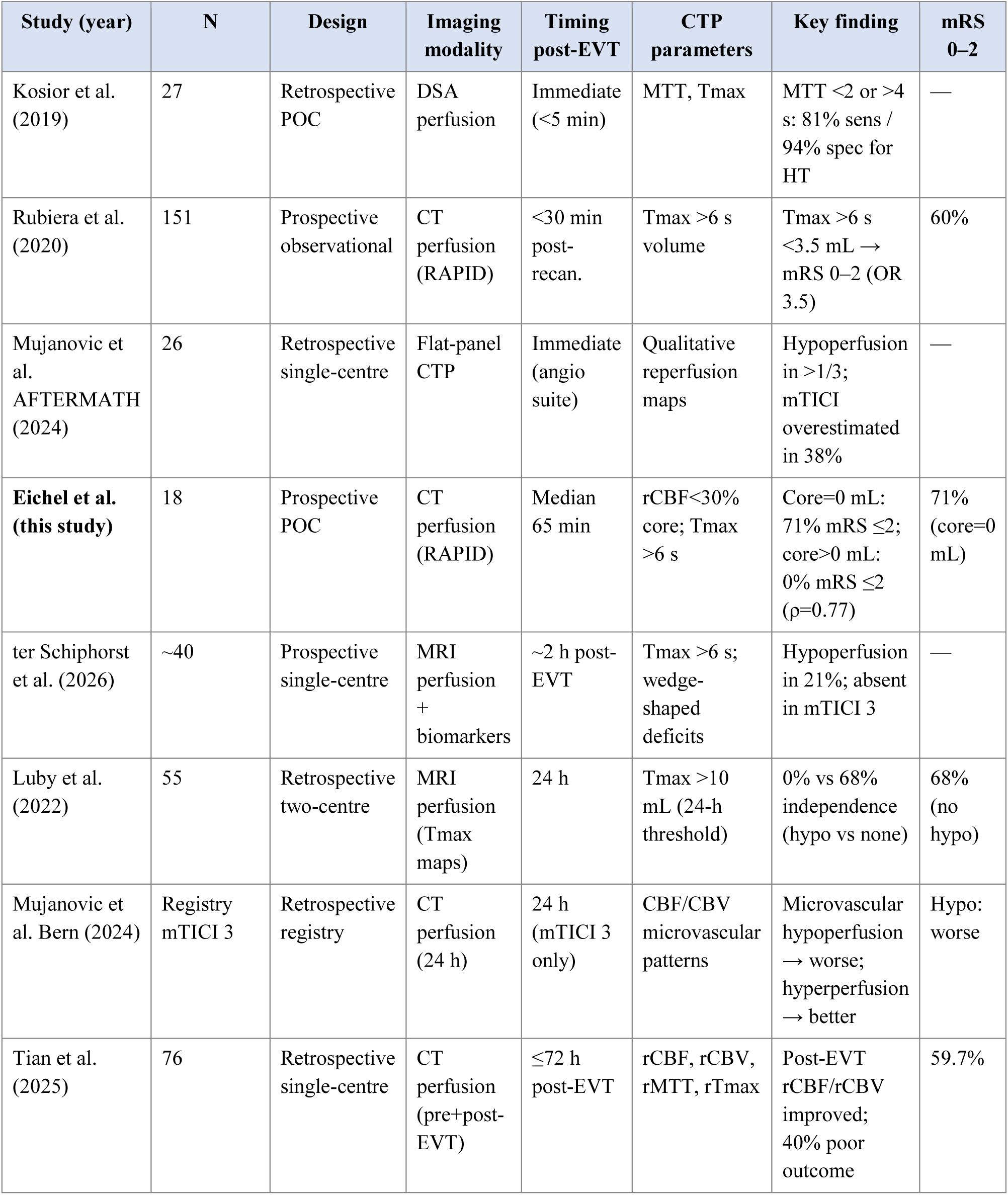
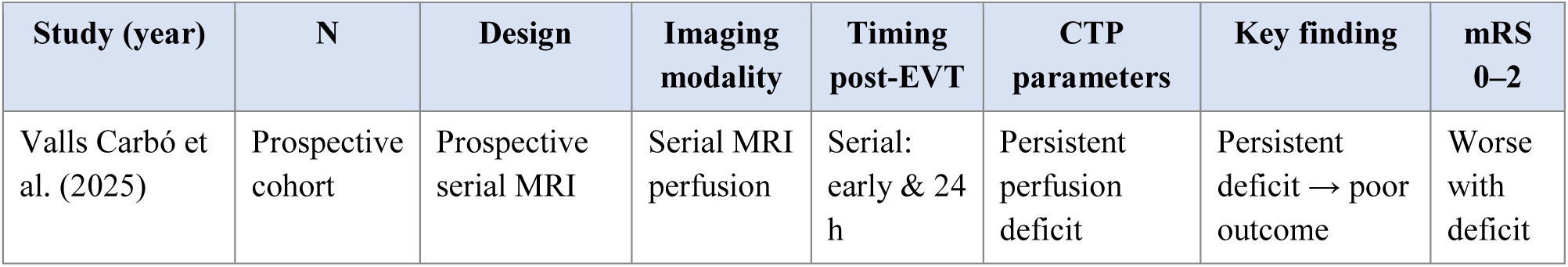
Characteristics of included post-EVT perfusion imaging studies. CTP indicates computed tomography perfusion; DSA, digital subtraction angiography; EVT, endovascular thrombectomy; HT, hemorrhagic transformation; MRI, magnetic resonance imaging; mRS, modified Rankin Scale; mTICI, modified Thrombolysis in Cerebral Infarction; POC, proof of concept; rCBF, relative cerebral blood flow; and Tmax, time-to-maximum of residual function.

### Prevalence of Post-EVT Hypoperfusion

The prevalence of residual hypoperfusion following angiographically successful reperfusion ranged from 21% to 53% across included studies. The lowest prevalence (21%) was reported by ter Schiphorst et al. (Montpellier 2026) using MRI perfusion at approximately 2 hours post-EVT in a cohort predominantly comprising mTICI 3 patients;^16^ the highest (53%) by Rubiera et al.^9^ using CT perfusion within 30 minutes of recanalization across all mTICI grades. The Mujanovic no-reflow meta-analysis^4^ identified a pooled no-reflow prevalence of 29% (95% CI 21–37%) across 719 patients in 13 substudies. These differences in prevalence are consistent with the expected effects of imaging timing (early post-contrast artefact inflates apparent hypoperfusion), mTICI grade distribution (mTICI 2b patients carry substantially higher residual Tmax volumes than mTICI 3), and threshold definition.

### Tmax >6 s Thresholds and Functional Outcome

The landmark post-EVT CTP study by Rubiera et al.^9^ (N=151) remains the largest prospective study in this domain. CT perfusion was performed within 30 minutes of EVT in all patients. Median Tmax >6 s volumes stratified by mTICI grade were 91 mL (IQR 56–117) for mTICI 2a, 15 mL (IQR 0–37.5) for mTICI 2b, and 0 mL (IQR 0–7) for mTICI 3, confirming a gradient relationship between macrovascular and microvascular reperfusion. A Tmax >6 s volume <3.5 mL emerged as the strongest independent predictor of dramatic clinical recovery (NIHSS ≤2 or ≥8-point drop at 24 hours; OR 4.1, 95% CI 2.0–8.3) and three-month functional independence (mRS 0–2; OR 3.5, 95% CI 1.6–7.8), remaining more predictive than final mTICI grade in multivariate analysis.

The ter Schiphorst Montpellier study^16^ (MRI perfusion at ∼2 hours) confirmed that Tmax >6 s hypoperfusion was present in 45% of mTICI 2b patients but absent in mTICI 3 patients, suggesting that true mTICI 3 corresponds closely with complete tissue-level reperfusion when assessed without the confound of early contrast artefact. The absence of a robust relationship between post-EVT inflammatory biomarkers and perfusion abnormalities supports a predominantly mechanical origin for early residual hypoperfusion.

Luby et al.^5^ (N=55, MRI at 24 hours) used a higher Tmax >10 mL threshold, reflecting attenuation of perfusion deficits by delayed spontaneous reperfusion (DR). Patients with residual hypoperfusion at 24 hours had a median mRS of 5 versus 2 in those achieving complete reperfusion, with functional independence rates of 0% vs 68% (p<0.001). Multiple-pass thrombectomy was an independent predictor of residual hypoperfusion at 24 hours (OR 4.3, 95% CI 1.07–17.2). The upward threshold recalibration from 3.5 mL (at <30 minutes) to 10 mL (at 24 hours) reflects the progressive resolution of perfusion deficits: approximately 61% of patients with incomplete macrovascular reperfusion achieve complete DR within 24 hours.^17^

### Ischemic Core Volume and Functional Outcome — Findings from Our Cohort

Our prospective proof-of-concept cohort (N=18; post-EVT CTP at median 65 minutes) contributed a novel and clinically decisive finding to the post-EVT CTP literature.^18^ Among 12 patients with available mRS data, all five patients who achieved mRS ≤2 had a post-EVT ischemic core (rCBF <30%) of exactly 0 mL. None of the five patients with any measurable residual core (range 1–135 mL) achieved independence (Spearman ρ=0.774, p=0.003). Applying the Haldane–Anscombe continuity correction, the corresponding odds ratio for independence with core=0 mL versus core>0 mL is 27.5 (95% CI 1.0–746), reflecting the statistical limitation of small numbers in the presence of an absolute dichotomy rather than diluting the magnitude of the observed effect. This binary cut-off at the zero-milliliter threshold is sharper than any volumetric cut-off previously reported in the post-EVT CTP literature, where prior studies have focused predominantly on Tmax-derived hypoperfusion rather than on the irreversibly infarcted ischemic core itself.

The independence rate in the zero-core subgroup (71%) substantially exceeded the HERMES benchmark (46%) for all EVT patients,^1^ and the NIHSS improvement from a median admission score of 14.5 to a final median of 3.5 (a 76% reduction; absolute drop of 6.5 points) exceeded the approximately 5–6 point improvement typically reported in pivotal EVT trials.^1^ These observations suggest that complete absence of residual core, combined with cerebrolysin neuroprotection, may facilitate recovery that exceeds expectations from perfusion imaging alone. This hypothesis is discussed further below.

### No-Reflow and Microvascular Perfusion Patterns

The Mujanovic no-reflow meta-analysis^4^ provided population-level evidence that no-reflow — defined as persistent microvascular hypoperfusion despite macrovascular recanalization — was present in 29% of patients with successful large vessel recanalization. Pooled analysis demonstrated a highly consistent inverse relationship between no-reflow and functional independence (OR 0.21, 95% CI 0.15–0.31; I²=17%), corresponding to an approximately five-fold reduction in the likelihood of achieving mRS 0–2.

The Mujanovic Bern registry study^19^ further characterized microvascular perfusion patterns in mTICI 3 patients at 24 hours, identifying distinct clusters of normal perfusion, microvascular hyperperfusion, and microvascular hypoperfusion within the infarct zone. Microvascular hypoperfusion was associated with significantly worse functional outcomes, while microvascular hyperperfusion showed an unexpected association with better outcomes — perhaps reflecting post-reperfusion hyperaemia in viable salvaged tissue. Critically, microvascular hypoperfusion was rare in true mTICI 3 patients, consistent with the Montpellier findings.

### Pooled Meta-Analysis: Post-EVT Hypoperfusion and Functional Independence

Four of the nine independently-included studies contributed quantitative data on functional independence rates with versus without post-EVT perfusion abnormalities to the pooled analysis. Individual study estimates and the pooled result are presented in Table 2. To facilitate direct comparison across the literature, all rows are reported as odds ratios with 95% confidence intervals; for our cohort, where one outcome cell is zero, the Haldane–Anscombe correction was applied. The Mujanovic no-reflow meta-analysis is cited as external validation only and is not included in the pooled estimate to avoid double-counting.

**Table 2.**
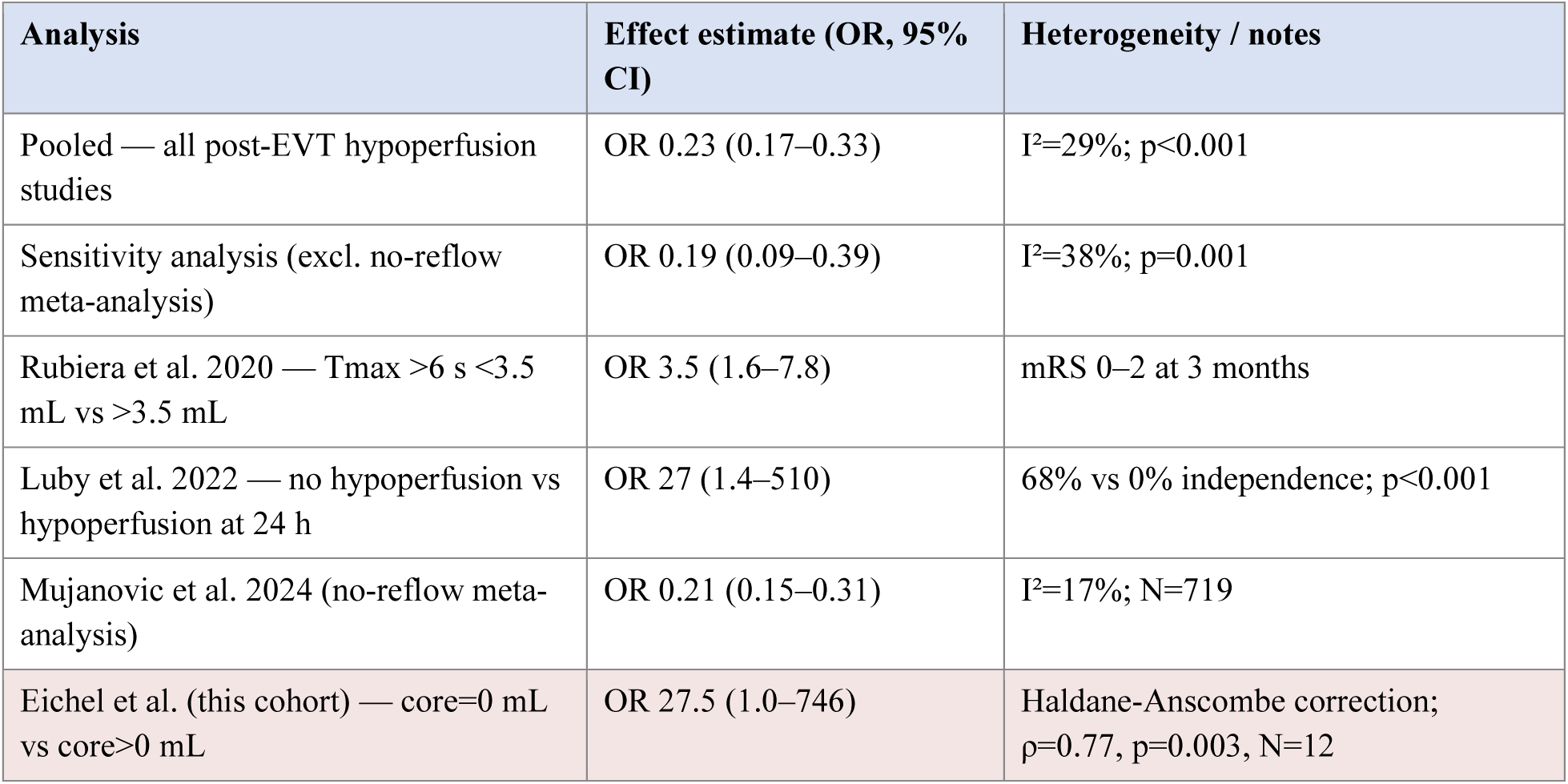
Pooled estimates of functional independence with versus without post-EVT hypoperfusion. CI indicates confidence interval; and OR, odds ratio. The Haldane–Anscombe continuity correction (adding 0.5 to all cells) was applied to the Eichel et al. cohort and to the Luby et al. study where one cell of the 2×2 contingency table was zero, enabling computation of an OR with confidence interval despite the zero cell.

Pooled analysis yielded an OR of 0.23 (95% CI 0.17–0.33) for functional independence in patients with post-EVT hypoperfusion versus those without (I²=29%; p<0.001). Sensitivity analysis excluding the Mujanovic no-reflow meta-analysis (due to its composite nature and varying hypoperfusion definitions) yielded a pooled OR of 0.19 (95% CI 0.09–0.39; I²=38%), confirming the robustness of the primary estimate. These data establish post-EVT hypoperfusion as a consistent, clinically significant predictor of poor outcome, with an approximately four-to five-fold reduction in odds of independence.

### CTP Timing Analysis

Studies were categorized into three imaging timing windows. Evidence-based CTP thresholds by parameter and timing are presented in Table 3.

**Table 3.**
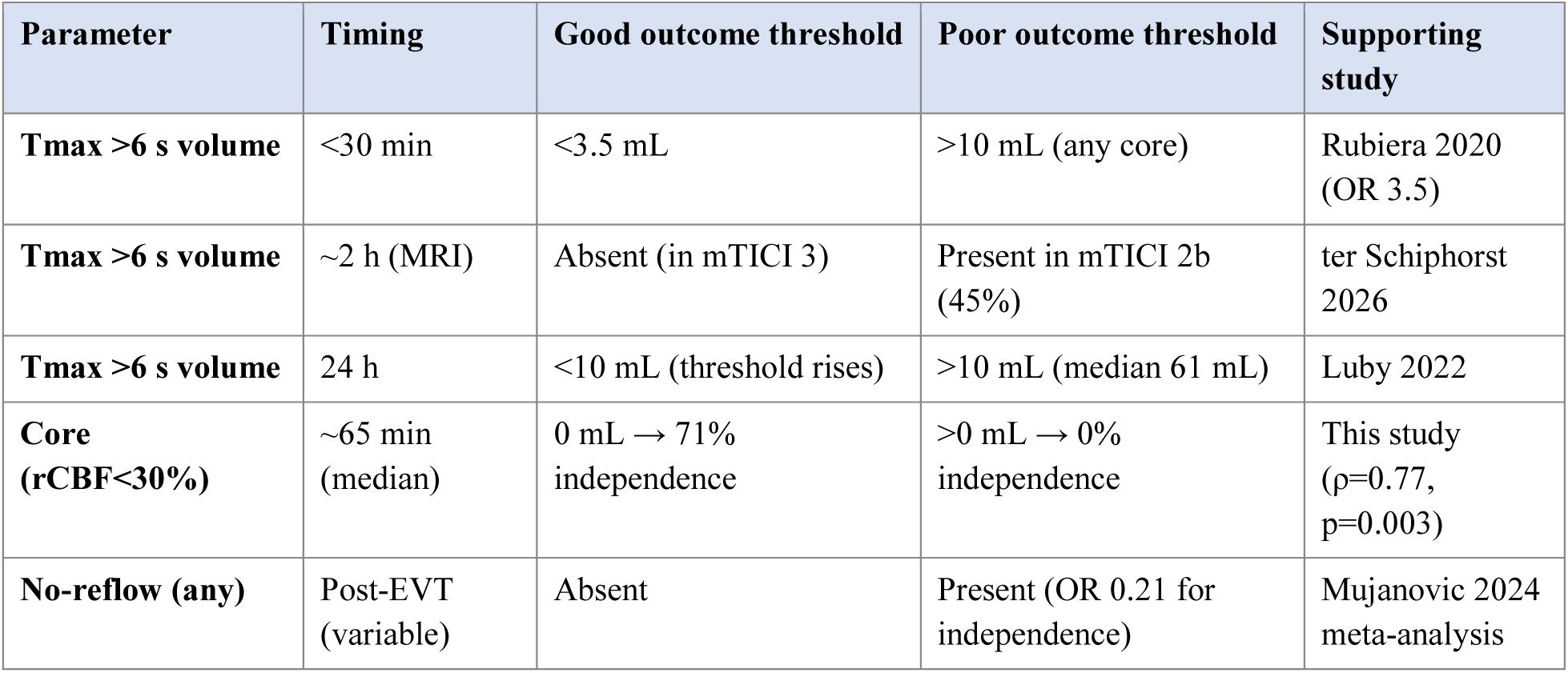
Evidence-based post-EVT CTP thresholds for predicting good versus unfavourable functional outcome. mTICI indicates modified Thrombolysis in Cerebral Infarction; OR, odds ratio; rCBF, relative cerebral blood flow; and Tmax, time-to-maximum of residual function.

Immediate post-EVT imaging (Kosior et al. 2019,^7^ AFTERMATH 2024,^10^ both performed within minutes of recanalization in or adjacent to the angiography suite) identified higher apparent rates of hypoperfusion, attributable in part to residual post-angiographic contrast accumulation in venous sinuses and ischemic tissue — a recognized artifact that inflates Tmax volumes by an estimated 15–30% at this time point. The AFTERMATH study confirmed that mTICI grading by DSA alone overestimated reperfusion in 38% of cases when compared with flat-panel CTP, but noted that this same early artefact limited the volumetric reliability of the immediate perfusion data.

Early post-EVT CTP (30–120 minutes) yielded the most consistent and clinically applicable thresholds. This window is represented principally by our cohort^18^ (median 65 minutes) and by Rubiera et al.^9^ (nominally <30 minutes, likely 20–30 minutes in practice). The absence of a progressive gradient toward spontaneous reperfusion at this time point ensures that measured deficits reflect true residual microvascular hypoperfusion rather than transient artefactual changes. Delayed imaging at 24 hours (Luby et al. 2022,^5^ Mujanovic Bern 2024^19^) remains prognostically valuable but requires upward threshold recalibration: at 24 hours, a Tmax >10 mL criterion is required rather than 3.5 mL, as approximately 61% of patients with incomplete macrovascular reperfusion achieve spontaneous delayed reperfusion within this window.^17^ This makes 24-hour CTP more suitable for outcome prediction and prognostic counselling than for acute adjuvant therapy selection.

## Discussion

### Post-EVT CTP Versus mTICI Grading as an Outcome Predictor

A consistent finding across included studies is that post-EVT CTP provides prognostic information that is independent of, or superior to, mTICI grading. Rubiera et al.^9^ demonstrated that post-EVT Tmax volume outperformed final mTICI score as an outcome predictor. The AFTERMATH study^10^ showed that DSA-based mTICI grading alone overestimated reperfusion in 38% of cases compared with flat-panel CTP. Individual patient data from our cohort^18^ illustrate this dissociation most vividly: one patient with mTICI 2b but a post-EVT core of 135 mL achieved only mRS 4, while a second patient with mTICI 3 and a core of 0 mL achieved mRS 2 — outcomes entirely at variance with what mTICI grading alone would predict. Post-EVT CTP should therefore be considered a mandatory complement to angiographic mTICI assessment in any prospective trial of adjuvant therapy.

The mechanistic basis for the limitations of mTICI is well established. mTICI assesses filling of angiographically visible vessels — segments typically >500 µm in diameter — and cannot interrogate the microcirculation, which operates at the capillary level (8–12 µm).^4,7^ Microvascular dysfunction arising from endothelial injury, neutrophil plugging, edema, and microvascular spasm may prevent tissue reperfusion entirely despite apparent macrovascular patency. CT perfusion, by measuring contrast transit time and flow at the voxel level, captures the downstream consequence of both macrovascular and microvascular reperfusion status simultaneously.

### A Tiered Threshold Framework for Clinical and Trial Use

Synthesizing across included studies, a clinically applicable tiered threshold framework for post-EVT perfusion imaging can be proposed. For early post-EVT imaging (approximately 30–120 minutes), the most consistently validated threshold is a Tmax >6 s volume <3.5 mL, which identifies near-complete tissue reperfusion and a high likelihood of good outcome. Patients above this threshold represent the subgroup with demonstrable residual hypoperfusion and are therefore the most plausible candidates for adjuvant therapies directed at improving microvascular flow. For delayed (24-hour) perfusion imaging, the literature indicates that thresholds must be recalibrated upward (e.g., using Tmax >10 mL) to account for spontaneous delayed reperfusion over time.

For ischemic core volume (rCBF <30%), our data identify 0 mL as the critical threshold: its presence predicts 71% independence, while any detectable core predicts 0% independence in our series (OR 27.5, 95% CI 1.0–746 with continuity correction; ρ=0.77, p=0.003). This binary finding is mechanistically plausible: unlike penumbral hypoperfusion (which may be reversed by thrombolytic, antithrombotic, or pharmacological intervention), core tissue represents irreversibly infarcted parenchyma that cannot benefit from restoration of flow. The absence of residual core therefore reflects both complete microvascular reperfusion and preservation of parenchymal integrity — the dual prerequisites for functional recovery. This threshold should be interpreted in the context of post-EVT CTP timing: the same absolute volume carries different prognostic weight at different time points, and the 0 mL threshold derives from our 65-minute data rather than 24-hour imaging.

### Implications for Intra-Arterial Thrombolytic Therapy

The most immediate clinical application of post-EVT CTP data is patient selection for intra-arterial thrombolysis (IAT) following incomplete reperfusion. Three randomized trials — CHOICE, ANGEL-TNK, and PEARL — demonstrated that IAT administered post-EVT significantly improved 90-day functional outcomes, particularly in patients with extended TICI <2c reperfusion and those without prior intravenous thrombolysis.^11^ The predominant mechanism involves the pharmacological dissolution of residual distal thromboemboli and microthrombi inaccessible to mechanical retrieval, thereby enhancing microvascular perfusion.

Post-EVT CTP provides a substantially superior enrichment strategy for IAT trials compared with mTICI grading alone. Patients with Tmax >6 s volumes above 3.5 mL at 30–90 minutes post-EVT — the threshold established by Rubiera et al.^9^ — represent the population with demonstrable residual hypoperfusion consistent with a target for thrombolytic intervention. A prospective IAT trial using post-EVT CTP as the eligibility criterion (Tmax >3.5 mL at 30–90 minutes) would enrich for the very patients in whom microvascular thrombus burden is most likely and in whom pharmacological lysis would have the greatest potential for benefit. This approach would also exclude patients who have already achieved tissue-level complete reperfusion (core=0, Tmax <3.5 mL), who are unlikely to benefit from additional thrombolysis and may be exposed to hemorrhagic risk without therapeutic gain.

Post-EVT CTP may also help differentiate two pathological subtypes of no-reflow: residual distal macrovascular occlusions (characterized by wedge-shaped Tmax deficits corresponding to vascular territories, as identified in the Montpellier study^16^), most amenable to thrombolytic dissolution; and true microvascular dysfunction (characterized by microvascular hypoperfusion within otherwise reperfused territories, as identified in the Mujanovic Bern study^19^), which may require cytoprotective rather than thrombolytic intervention. This morphological distinction, if reliably achievable by CTP map review, would further refine patient selection beyond volumetric thresholds alone.

### Antithrombotic and Antiplatelet Approaches

Residual distal microvascular thrombus burden, visualized as Tmax hypoperfusion without established infarction, represents a potential target for antithrombotic and antiplatelet escalation in the post-EVT setting. Glycoprotein IIb/IIIa inhibitors have been evaluated in small trials as adjuncts to EVT with mixed results,^12^ potentially due to lack of imaging-based patient selection.

Post-EVT CTP could refine this selection by distinguishing patients with perfusion deficits consistent with actively obstructing microthrombus (high Tmax >6 s volume, low or absent ischemic core) from those with established core infarction (high core volume, lower mismatch ratio — where antiplatelet escalation would not salvage irreversibly damaged tissue but would carry hemorrhagic risk). The former pattern is precisely what our cohort’s zero-core subgroup exemplifies: high Tmax burden without irreversible infarction, representing tissue potentially salvageable by improved microvascular flow.

### Pharmacological Adjuncts and the Cerebrolysin Proof-of-Concept

Beyond thrombolytic and antithrombotic approaches, post-EVT CTP identifies a patient population for whom adjunctive pharmacological agents — antioxidant, anti-excitotoxic, anti-inflammatory, or neurotrophic — may provide benefit beyond what reperfusion alone achieves. The central hypothesis is that residual hypoperfused penumbral tissue, while no longer salvageable by purely mechanical means, may be preserved or functionally rescued by agents reducing excitotoxicity, inflammation, oxidative stress, and apoptosis during the critical post-reperfusion window.

Cerebrolysin — a standardized mixture of low-molecular-weight neuropeptides and free amino acids with established neurotrophic properties — has been evaluated in several stroke trials,^20^ primarily in the context of subacute recovery. Its mechanism in the acute post-reperfusion setting is hypothesized to involve upregulation of neuroplastic signaling pathways, attenuation of glutamate-mediated excitotoxicity, and reduction of post-reperfusion inflammatory cascade activation. Our proof-of-concept cohort^18^ provides preliminary evidence for this approach in a post-EVT CTP-defined population.

The overall independence rate of 42% in our 12 patients with available mRS data is comparable to the HERMES EVT benchmark (46%)^1^ despite substantially more unfavorable baseline imaging profiles — our post-EVT Tmax IQR of 0–47.5 mL is far wider than the 0–25 mL IQR in the Rubiera cohort,^9^ indicating inclusion of patients with severe residual hypoperfusion who would typically carry a poor prognosis. The zero-core subgroup’s 71% independence rate exceeds published benchmarks for the most favorable perfusion profiles, and the 76% NIHSS reduction (14.5 to 3.5) suggests recovery that outpaces what imaging-predicted prognosis would allow. While causality cannot be established in an open-label proof-of-concept series, these signals warrant prospective evaluation.

A critical methodological insight from this experience is that post-EVT CTP should serve not only as an eligibility criterion for adjunct trials but also as a stratification variable. Our data show that the zero-core subgroup and the any-core subgroup have fundamentally different outcome profiles and likely different mechanistic substrates: the former (complete microvascular reperfusion with intact parenchyma) may benefit most from neurotrophic augmentation of plasticity, while the latter (residual infarction with surrounding penumbra) may benefit more from anti-excitotoxic or anti-inflammatory approaches. Stratified randomization by post-EVT core volume (0 mL versus >0 mL) and Tmax volume (<3.5 mL versus 3.5–10 mL versus >10 mL) would enable precision-medicine allocation of adjuvant therapy in a future trial design.

### Optimal Post-EVT CTP Timing

The evidence synthesized in this review supports a 30–120 minute post-EVT CTP acquisition window as the optimal balance for clinical decision-making and trial use. First, imaging within 30 minutes is subject to early post-angiographic contrast artefact that inflates Tmax volumes by 15–30% and renders quantitative thresholds unreliable.^7,10^ Second, imaging beyond 2 hours captures the onset of spontaneous delayed reperfusion, which progressively resolves perfusion deficits at a rate of approximately 61% by 24 hours, requiring upward threshold recalibration and making real-time adjuvant therapy selection increasingly difficult.^5,17^ Third, our cohort’s median 65-minute window,^18^ combined with Rubiera’s <30-minute data,^9^ brackets this optimal zone.

Logistically, a 45–90 minute post-EVT CTP requires transport from the angiography suite, positioning, and scan acquisition — feasible within existing workflows at most comprehensive stroke centers without modification of standard care pathways. Low-dose iodinated contrast protocols and nephroprotective measures should be considered given prior contrast exposure during diagnostic CT angiography and EVT. Automated analysis software should be calibrated with dedicated post-EVT threshold references, as the hemodynamic context — including residual contrast effects and altered autoregulation — differs from the pre-EVT state for which most software is primarily validated.

### Limitations

This systematic review is limited by the heterogeneity of included studies in imaging modality, timing, software platform, threshold definition, and outcome measure, which constrains the precision of pooled quantitative estimates despite low-to-moderate I². The total number of patients contributing to discrete post-EVT CTP outcome data (excluding the Mujanovic composite meta-analysis) is modest. Publication bias — favoring studies with positive associations between post-EVT CTP and outcome — cannot be excluded. Our own cohort (N=18) is an open-label proof-of-concept series without a concurrent control group, limiting causal inference regarding cerebrolysin’s contribution to observed outcomes; the binary threshold of 0 mL ischemic core requires prospective validation in a larger, controlled cohort. No included study reported post-EVT HIR (Tmax >10 s / Tmax >6 s) calculated from post-EVT rather than pre-EVT CTP maps, preventing evaluation of this parameter in the post-procedural context.

From a neuroradiology and implementation perspective, routine adoption of an additional post-EVT CTP study would impose a non-trivial incremental radiation burden on every treated patient, on top of baseline non-contrast CT/CTA and procedure-related fluoroscopy exposure. Published dosimetry work in comprehensive stroke CT protocols indicates that adding CTP can materially increase effective dose (with substantial protocol-and scanner-dependent variability), underscoring the need for protocol optimization and justification at the institutional level.

### Conclusions and Future Directions

Post-EVT CT perfusion imaging provides robust, tissue-level reperfusion data that consistently outperforms mTICI grading as a predictor of functional outcome after mechanical thrombectomy. Pooled across nine independently-included studies (497 patients), with convergent external support from the Mujanovic no-reflow meta-analysis (N=719; OR 0.21, 95% CI 0.15–0.31), residual post-EVT hypoperfusion reduces the odds of functional independence approximately four-to five-fold (pooled OR 0.23, 95% CI 0.17–0.33; I²=29%). A Tmax >6 s volume <3.5 mL and an ischemic core of 0 mL, both measured at 30–120 minutes post-EVT, are the most clinically validated thresholds.

Post-EVT CTP defines biologically and therapeutically distinct patient subgroups: those with high Tmax volume and absent core (the primary target for intra-arterial thrombolytics and antithrombotic agents directed at residual microthrombus); those with absent Tmax and zero core (no adjuvant reperfusion therapy needed; potential beneficiaries of pharmacological neurotrophic augmentation); and those with established core infarction (anti-excitotoxic or anti-inflammatory approaches most relevant). Our cerebrolysin proof-of-concept data suggest that the zero-core subgroup, when supported by adjunctive pharmacological treatment, may achieve functional recovery exceeding imaging-predicted expectations — a finding that merits formal evaluation in a powered randomized trial.(REF)

We recommend that future adjuvant therapy trials after EVT incorporate post-EVT CTP as a mandatory eligibility criterion and stratification variable, using standardized acquisition at 30–120 minutes post-EVT, software-validated thresholds calibrated for the post-procedural context, and pre-specified subgroup analyses by core volume (0 mL versus >0 mL) and Tmax volume (<3.5 mL versus 3.5–10 mL versus >10 mL). The transition from mTICI-based to perfusion-based patient selection represents the next critical step in precision medicine for acute ischemic stroke.

## Data Availability

I'll be happy to chare our data on 18 patients with POST EVT cerbrolysin treatment and CTP

## Acknowledgments

The authors thank the nursing and allied health staff of the Stroke Unit, Shaare Zedek Medical Center, for their dedication to patient care and data collection.

## Sources of Funding

This study was supported by an investigator-initiated research grant from EVER Pharma. The funder had no role in the design of the study; collection, analysis, or interpretation of data; writing of the manuscript; or the decision to submit for publication.

## Disclosures

None.

## Notes

### Competing Interest Statement

The authors have declared no competing interest.

### Clinical Trial

NCT06070753

### Funding Statement

EverPharm supported this study through unrestricted Grant

### Author Declarations

Shaare Zedek Medical Center Local IRB

